# Longitudinal and Quantitative Fecal Shedding Dynamics of SARS-CoV-2, Pepper Mild Mottle Virus and CrAssphage

**DOI:** 10.1101/2023.02.02.23285391

**Authors:** Peter J. Arts, J. Daniel Kelly, Claire M. Midgley, Khamal Anglin, Scott Lu, Glen R. Abedi, Raul Andino, Kevin M. Bakker, Bryon Banman, Alexandria B. Boehm, Melissa Briggs-Hagen, Andrew F. Brouwer, Michelle C. Davidson, Marisa C. Eisenberg, Miguel Garcia-Knight, Sterling Knight, Michael J. Peluso, Jesus Pineda-Ramirez, Ruth Diaz Sanchez, Sharon Saydah, Michel Tassetto, Jeffrey N. Martin, Krista R. Wigginton

**Affiliations:** Department of Civil and Environmental Engineering, University of Michigan, Ann Arbor, MI, USA; Department of Epidemiology and Biostatistics, University of California, San Francisco, CA, USA; Institute for Global Health Sciences, University of California, San Francisco, CA, USA; Division of Hospital Medicine, UCSF, San Francisco, CA, USA; F.I. Proctor Foundation, University of California, San Francisco, CA, USA; National Center for Immunizations and Respiratory Diseases, Centers for Disease Control and Prevention, Atlanta, GA, USA; Department of Microbiology and Immunology, UCSF; Department of Epidemiology, University of Michigan, Ann Arbor, MI, USA; Department of Civil & Environmental Engineering, Stanford University, Stanford, CA, USA; School of Medicine, University of California, San Francisco, CA, USA; Division of HIV, Infectious Disease, and Global Medicine, UCSF, CA, USA

## Abstract

Wastewater-based epidemiology (WBE) emerged during the COVID-19 pandemic as a scalable and broadly applicable method for community-level monitoring of infectious disease burden, though the lack of high-quality, longitudinal fecal shedding data of SARS-CoV-2 and other viruses limits the interpretation and applicability of wastewater measurements. In this study, we present longitudinal, quantitative fecal shedding data for SARS-CoV-2 RNA, as well as the commonly used fecal indicators Pepper Mild Mottle Virus (PMMoV) RNA and crAss-like phage (crAssphage) DNA. The shedding trajectories from 48 SARS-CoV-2 infected individuals suggest a highly individualized, dynamic course of SARS-CoV-2 RNA fecal shedding, with individual measurements varying from below limit of detection to 2.79×10^6^ gene copies/mg - dry mass of stool (gc/mg-dw). Of individuals that contributed at least 3 samples covering a range of at least 15 of the first 30 days after initial acute symptom onset, 77.4% had at least one positive SARS-CoV-2 RNA stool sample measurement. We detected PMMoV RNA in at least one sample from all individuals and in 96% (352/367) of samples overall; and measured crAssphage DNA above detection limits in 80% (38/48) of individuals and 48% (179/371) of samples. Median shedding values for PMMoV and crAssphage nucleic acids were 1×10^5^ gc/mg-dw and 1.86×10^3^ gc/mg-dw, respectively. These results can be used to inform and build mechanistic models to significantly broaden the potential of WBE modeling and to provide more accurate insight into SARS-CoV-2 prevalence estimates.

## Introduction

SARS-CoV-2 RNA is commonly shed in the feces of individuals infected by the virus.^1–3^ This has led to the widespread adoption of Wastewater-Based Epidemiology (WBE) for tracking community levels of COVID-19. Measurable concentrations of SARS-CoV-2 RNA in wastewater correlate with measures of COVID-19 incidence,^4–8^ hospitalizations,^6,9,10^ and deaths.^8,11^ Although these correlations provide proxies for the relative levels of transmission in a sewershed over time, a mechanistic link between fecal shedding of SARS-CoV-2 RNA with wastewater monitoring would substantially strengthen the utility of WBE methods. For example, epidemiological models estimating total infections in a community (i.e. prevalence)^12,13^ or effective reproductive number (Re)^14^ rely on the integration of fecal shedding and wastewater transport models. Additionally, establishing a link to fecal shedding would help optimize the application and interpretation of WBE including identifying desirable sewershed sizes^15^ or sampling frequencies,^16^ knowing when and how to normalize measurements,^17^ or explaining causes of observed differences between wastewater and clinical trends.^8,10,18^

Mechanistic descriptions of community-wide WBE systems require accurate descriptions of fecal shedding, which are generated from longitudinal SARS-CoV-2 stool data that is both quantitative and externally-valid. Here, externally-valid describes data that features a basis of measurement which is useful in a wide variety of applications. To date, there have been numerous published studies on SARS-CoV-2 RNA prevalence and viral load in stool, including several reviews.^1–3,19–25^ Nearly all, however, lack the necessary information to be applicable for WBE models. For example, several studies reported a prevalence of fecal shedding in samples or individuals, but do not describe critical information including the window of time sampled, the coverage of sampling over this period, the precise times that samples were collected in the infection (e.g., days after initial symptom onset), or records of stool density or solid content. Additionally, quantitative data by qPCR is often reported in terms of cycle threshold rather than absolute abundance, or is missing methodological information recommended by *M*inimum *I*nformation for Publication of *Q*uantitative Real-Time PCR *E*xperiments (MIQE) guidelines (e.g., method detection limits, controls, etc.).^26,27^

CrAss-like phage (crAssphage) and pepper mild mottle virus (PMMoV) concentrations are often measured alongside SARS-CoV-2 nucleic acid concentrations in wastewater samples. The measured concentrations are used to normalize the pathogen viral nucleic acid measurements to account for differences in wastewater fecal strength and methodological viral recovery. To date, there is very little quantitative data available on the levels and temporal trends of these biomarkers in feces. This quantitative data is important for mechanistically linking normalized wastewater pathogen measurements with community disease burdens and also for identifying the scenarios in which normalizing by biomarker levels is appropriate. Indeed, models have been developed using PMMoV fecal shedding to estimate fecal strength of wastewater streams,^12,28^ though the available data of PMMoV in human feces is currently too limited to reliably use such a model.

In this study, we present quantitative fecal shedding trajectories of SARS-CoV-2 RNA and commonly used biomarkers PMMoV and crAssphage from 48 individuals who tested positive for COVID-19. The data collected include sufficient methodological information for their application in materials balance models, such as those associated with WBE and environmental risk assessment. The results show a highly individualized course of SARS-CoV-2 shedding over the first 30 days after initial onset of symptoms (ASO). Shedding of PMMoV and crAssphage was also highly variable between individuals over the same sampling period, albeit exhibiting different and distinct shedding patterns. Together, these results provide critical data for advancing the utility of WBE as a public health tool.

## Results

### Cohort Description

In total, 48 individuals provided stool samples for this study. Four of the 48 individuals did not experience symptoms of acute COVID-19. Of those with symptoms, the earliest stool samples were collected at three days pre-symptom onset and the latest samples were collected on day 28 after initial acute symptom onset. 382 samples were collected from 26 index and 22 household contacts that became infected, between September 2020 and April 2021. The demographics of the cases were: 58% female, 42% male; and 14.6% 0-17 years old, 70.8% 18-55 years old, and 14.6% above 55 (additional information in Table S1). The ethnicity/race break-down was as follows: 23% Hispanic/Latino, 53% white, 3% Black/African American, 13% Asian, and 3% Other race/ethnicity. The cohort also contained 4 individuals who were fully vaccinated for SARS-CoV-2.

### SARS-CoV-2 Fecal Shedding

#### Analysis Summary

Quantitative measurements of SARS-CoV-2 RNA were conducted on 382 samples from 48 individuals (Figure 1A). The median number of samples collected per individual was 9, with a range of 1 to 15 samples per individual. Samples were collected between -3 to 28 days ASO. Assays targeting the viral genes N and ORF1a exhibited a log-linear correlation (r^2^ = 0.85, figure S1A); therefore, all subsequent analyses of SARS-CoV-2 RNA shedding used only the quantitative N gene data. A volume of bovine coronavirus modified live-virus vaccine (BCoV) was spiked in each sample prior to extraction and monitored to ensure sufficient recovery through processing. Three samples were ultimately excluded from analysis due to BCoV recoveries under 50%, and thus our final dataset consisted of 379 samples. The limit of blank for our SARS-CoV-2 RNA concentrations, determined as the upper 95% confidence limit of the negative extraction control,^29^ ranged from 11.2 to 1550 gc/mg-dw, with differences between analytical runs and between samples due to varying levels of background in our negative extraction controls and differences in the sample stool percent solid values. The limit of blank (LOB) was used as the threshold of positivity for ddPCR to account for run-to-run changes in background levels of amplification. The method of calculation for the LOB is included in the ddPCR methods section.

**Figure 1:**
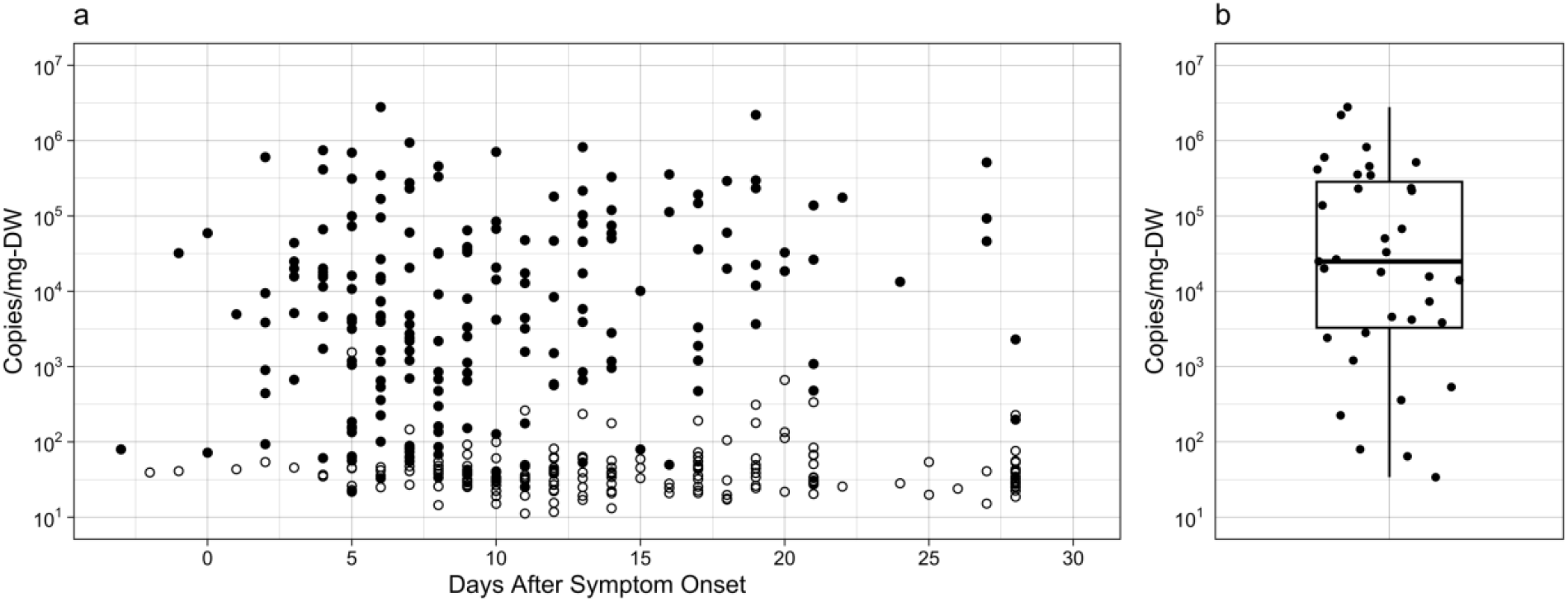
SARS-CoV-2 RNA fecal shedding data A) Longitudinal plot of all SARS-CoV-2 N gene measurements. Empty symbols represent sample measurements below the limit of blank. B) Boxplot and individual data points summarizing the peak fecal shedding magnitude for each individual with at least one sample above the limit of blank.

#### Shedding prevalence among population

In summary, 50.9% (193/379) of all collected samples resulted in SARS-CoV-2 N gene measurements above the limit of blank, and 72.9% (35/48) of the individuals contributed at least one sample above the limit of blank. Of the remaining 27.1% of all individuals who had no stool samples positive for SARS-CoV-2 RNA, 3 individuals (4542, 4567, and 4571) provided fewer than three samples over the course of the sampling period, limiting the coverage of a potential period of fecal shedding (Figure S2). Here, if we only include individuals who contributed at least three samples spanning at least 15 days between the earliest and latest collected sample,the proportion of participants without positive measurements decreased to 22.6% (7/31). It remains possible that these individuals excreted SARS-CoV-2 RNA on days when samples were not collected, or at levels that were below our LOB. Three of the four vaccinated individuals included in this cohort had measurable SARS-CoV-2 RNA in their stool at some point in the sampling period (Supplementary Data).

#### Shedding prevalence over time

To understand the prevalence of fecal shedding at each day after initial acute onset of symptoms in the studied population, we calculated the proportion of samples collected for a specific day that were positive for SARS-CoV-2 RNA (Figure 2A). Prior to symptom onset, as well as the first day ASO, the numbers of samples per day are limited (Figure 2B), making it difficult to draw conclusions about the prevalence of fecal shedding during this period. We note that two of the four samples collected pre-symptom onset were positive for SARS-CoV-2 RNA, corresponding to two of three individuals that contributed pre-symptomatic samples. These results provide important evidence of pre-symptomatic fecal shedding of SARS-CoV-2 RNA. Our sample set includes more samples per day from day 2 ASO through day 20 ASO, and thus presents a clearer picture of shedding prevalence during that time. The prevalence of fecal shedding peaks at 85.7% on days two and 3 ASO, and then this percentage decreases until reaching 10% on day 28 ASO.

**Figure 2:**
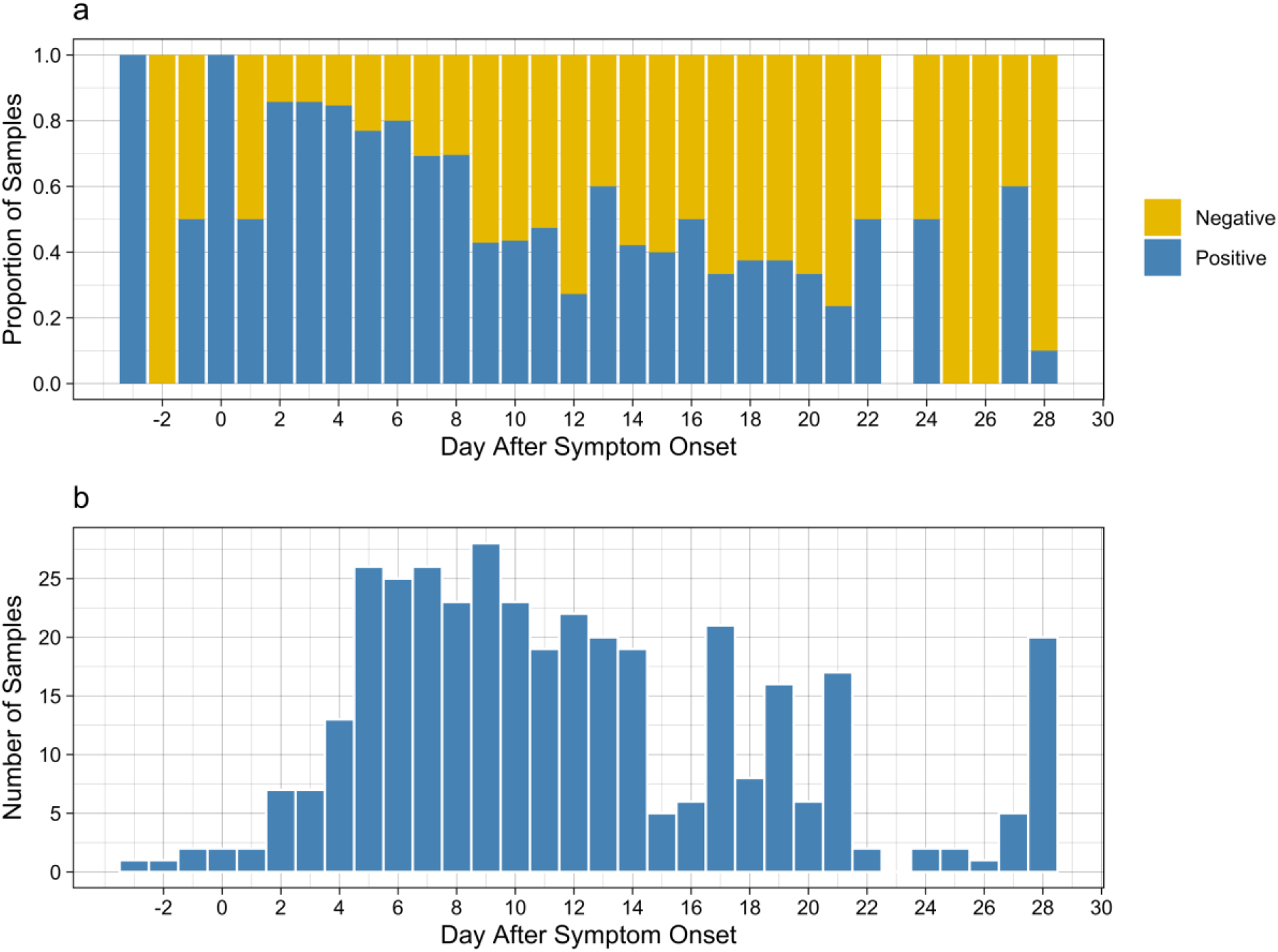
A) The proportion of stool samples on each day after symptom onset where SARS-CoV-2 N gene was measured above the limit of blank. B) A histogram showing the number of samples measured on each day after symptom onset.

A mixed effect logistic regression was applied to the shedding prevalence data for each day to test the fixed effects of day ASO and sex, with the influence of each individual as the random effects (Table S3). Our results suggest that the decrease in probability of SARS-CoV-2 fecal shedding with increasing days ASO with an odds ratio of 0.597 (log10 estimate = -0.22933), indicating a decrease in likelihood of shedding with each additional day. The effect of sex on the observed fecal shedding was not statistically significant. Demographics such as age, symptomaticity, vaccination status, and ethnicity were excluded due to insufficient sample sizes across groups to provide significant information.

#### Quantitative measurements

The quantitative RT-ddPCR measurements coupled with stool percent solids measurements provide externally valid SARS-CoV-2 RNA absolute abundance data. The geometric mean of all measurements above the LOB was 5.25×10^3^ gc/mg-dw, the geometric standard deviation was 17.9 gc/mg-dw, and the median was 4.83×10^3^ gc/mg-dw. The maximum shedding value observed across all 379 samples was 2.79×10^6^ gc/mg-dw and the minimum positive shedding value observed was 21.7 copies/mg-dw. The peak SARS-CoV-2 N gene concentration for individuals with sufficient sample coverage varied over approximately 6 orders of magnitude (Figure 1B). The maximum and minimum measured peak values for individuals were 2.79×10^6^ gc/mg-dw and 33.9 gc/mg-dw, respectively. The median peak shedding value was 1.91×10^4^ gc/mg-dw, the geometric mean was 1.27×10^4^ gc/mg-dw, and the geometric standard deviation was 17.9 gc/mg-dw.

### PMMoV and CrAssphage Fecal Shedding

#### Analysis Summary

PMMoV and crAssphage nucleic acids were measured in each stool sample to observe the biological variability of commonly used fecal biomarkers between individuals, and within individuals over the sampling period (Figure 3). PMMoV RNA was measured by ddPCR on the same day as the SARS-CoV-2 and BCoV RNA measurements, and the 3 samples with BCoV recoveries less than 50% were excluded from the data analyses. An additional 12 samples were excluded from PMMoV data analysis due to a lack of sufficient separation between background and positive droplets. This was observed in isolated samples from 4 individuals (4512, 4514, 4545, 4576), and for every sample from one individual (4577). This result could be due to the level degeneracy between the designed assay target and the actual sequences found in the samples.

**Figure 3:**
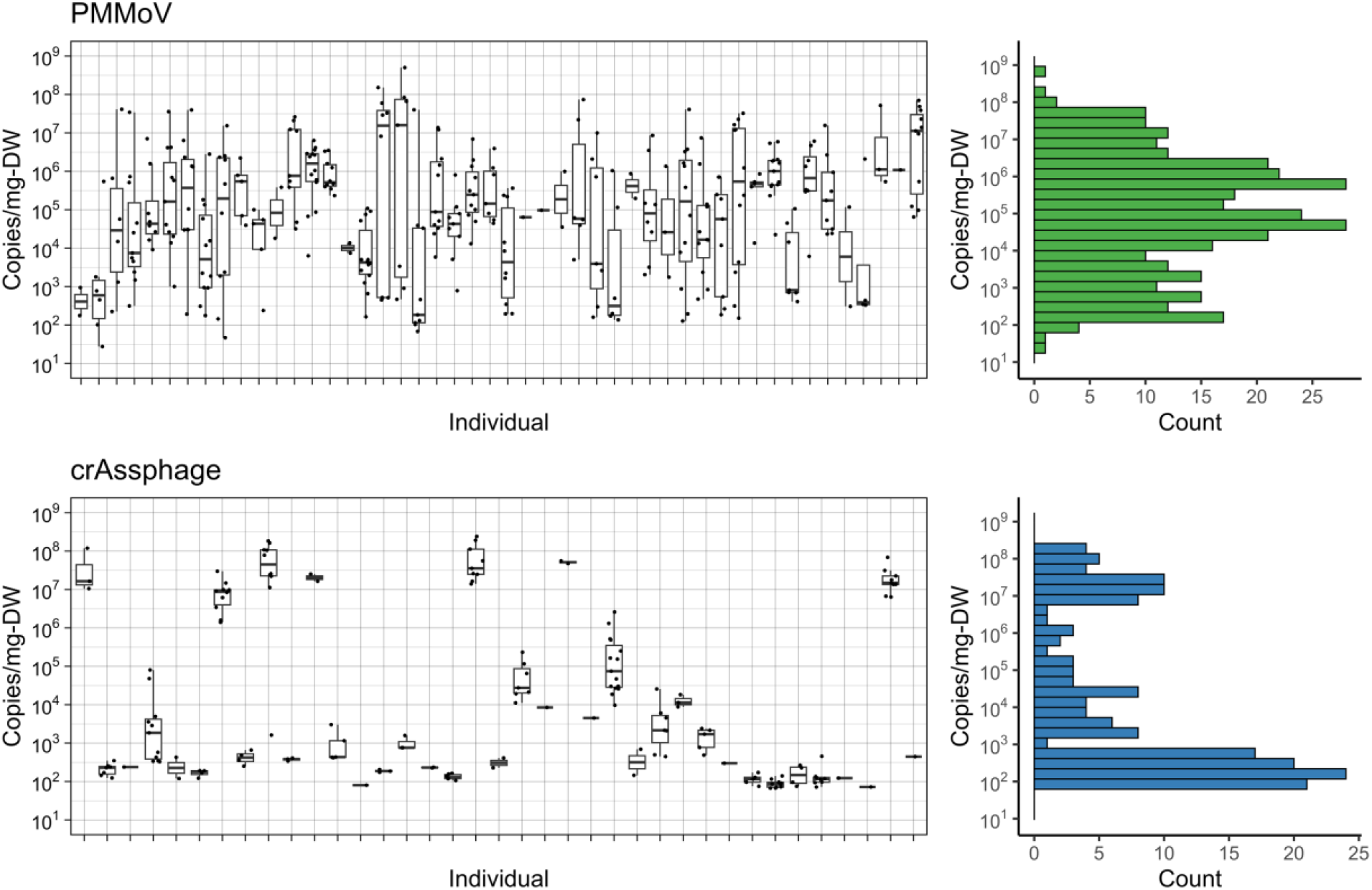
Above LOB or LOD shedding measurements and histogram of PMMoV (top) and crAssphage (bottom) for all samples in this study, as measured by qPCR (crAssphage) or RT-ddPCR (PMMoV). In the box plots (left), each box summarizes all measurements available for an individual with the bar indicating the median shedding value. The histograms (right), summarize the distribution of shedding magnitude for the entire set of above detection limit measurements.

#### Pepper mild mottle virus (PMMoV)

For PMMoV, 95.9% (352/367) of the measured samples had values above the LOB (mean LOB = 397 gc/mg-dw) and 100% (48/48) of individuals had at least one sample above the LOB. Longitudinal trajectories of PMMoV shedding for each individual are included in figure S3 The median concentration of PMMoV RNA in feces was 1.00×10^5^ gc/mg-dw and the maximum concentration observed was 5.0×10^8^ gc/mg-dw. PMMoV RNA measurements varied highly between individuals and over the sampling period. Some participants had consistent concentrations of PMMoV across many stool samples collected, whereas other donors had concentrations that varied by orders of magnitude between samples (Figure 3). The median range of measurable PMMoV RNA in individuals with at least 3 samples was 6.98×10^6^ gc/mg-dw.

#### CrAss-like phage (crAssphage)

CrAssphage DNA shedding was observed less frequently than PMMoV RNA shedding in the study participants. Longitudinal trajectories of crAssphage shedding for each individual are included in figure S4. In summary, 48.2% (179/371) of all samples were above the assay limit of detection (LOD) (mean LOD = 25.3 gc/mg-dw) and 79% (38/48) of individuals had at least one sample above the LOD. For qPCR, an assay limit of detection was used as the threshold of positivity, the method of LOD calculation is included in the qPCR methods section. Of the samples that were positive for crAssphage DNA, the maximum fecal concentration was 2.41×10^8^ gc/mg-dw and the median concentration was 2.13×10^3^ gc/mg-dw. CrAssphage DNA in samples from some individuals were consistently high, while others consistently shed much lower concentrations or no detectable crAssphage DNA. CrAssphage DNA fecal shedding was more consistent for individuals over time than PMMoV RNA shedding. The median range of shedding values within an individual was 368 gc/mg-dw for crAssphage, compared with 6.98×10^6^ gc/mg-dw for PMMoV. For each individual, crAssphage DNA levels varied less than three orders of magnitude, while for PMMoV RNA, the range of concentrations shed by each individual was greater than three orders of magnitude in 44 out of the 45 individuals with at least two samples. Interestingly, crAssphage concentrations in feces exhibited a bimodal distribution with most positive individuals shedding below 10^4^ gc/mg-dw (25/38) and a smaller fraction of individuals shedding above 10^6^ gc/mg-dw (7/38). An even smaller fraction of individuals exhibit intermediate magnitudes of shedding between 10^4^ and 10^6^ gc/mg-dw (6/38).

A mixed effect linear regression was applied to each set of shedding data to observe differences in the concentration of each biomarker shed in feces with fixed effect variables of sex, SARS-CoV-2 fecal shedding, and the concentration of the other biomarker shed. The output variable for these analyses was log10(gc/mg-dw) .The estimates and associated statistics associated with these results are included in table S4. Individual differences were investigated as the random effect. In the case of both crAssphage and PMMoV, none of the fixed effects had a statistically significant effect on the biomarker being shed. The random effects associated with PMMoV shedding were less variable than those associated with crAssphage, summarized by standard deviations of 0.824 and 2.164 log_10_(gc/mg-DW), respectively; reflecting the observations discussed above and shown in figure 3.

## Discussion

Here, we report quantitative, and externally valid data for fecal shedding of several targets of interest for WBE in SARS-CoV-2 infected individuals. This study features a large sample size and good sampling coverage over approximately 30 days. The resulting novel dataset facilitates important observations about the presence, magnitudes, and trends of viral nucleic acid fecal shedding amongst individuals. We report evidence of fecal shedding of SARS-CoV-2 RNA among a few pre-symptomatic, vaccinated individuals

There has been a sustained interest in the fraction of infected individuals that shed SARS-CoV-2 in feces, and we observed that approximately 22% of individuals that provided multiple samples did not shed SARS-CoV-2 in their feces up to day 28 ASO. A recent meta-analysis of fecal shedding of 38 individuals saw 51.9% of individuals did not shed SARS-CoV-2 RNA in feces; inspection of the included studies, however, the median number of samples collected per individual was 2.^3^ Presumably, if more samples were collected from each study participant, this percentage may decrease. The observed 22% percent, reported in our study, included only those individuals with > 3 samples collected over > 15 days of their infection; it is likely that increasing the resolution of samples collected over the first 30 days of infection (e.g., daily) would result in an even lower percentage of individuals without positive SARS-CoV-2 samples.

Our high-resolution data and relatively large study population provided a unique description of the prevalence of shedding through the first 30 days ASO. Approximately 80% of samples collected within the first five days were positive for SARS-CoV-2, this percentage dropped to 10% of samples at 28 days ASO. Natarajan and colleagues (2021)^2^ collected samples from a large number of individuals (120) but collected only three samples per individual over the 30 days ASO. They found that approximately 75% of individuals were shedding SARS-CoV-2 RNA in samples collected between days 0 and 7 and less than 25% of individuals were shedding SARS-CoV-2 RNA in samples collected between days 22 and 35 ASO. Although our data set ends at 28 days ASO, Natarajan et al. also observed shedding in a small fraction of individuals up to 288 days ASO.

The quantitative SARS-CoV-2 data presented here highlights the large variability in fecal shedding trajectories and magnitudes of SARS-CoV-2 between individuals. For example, of those that shed SARS-CoV-2 RNA at some point during their infection, peak shedding varied from as soon as 2 days ASO to 27 days sampling period (Figure S2) and peak values span approximately 4 orders of magnitude (Figure 1B). A study by R. Wölfel and colleagues (2020)^1^ contains the most comprehensive fecal shedding trajectories to date, with a set that included 60 stool samples from 9 participants with mild to moderate COVID-19. Wölfel et al. also observed peak shedding values that ranged 4 orders of magnitude. Since they reported RNA copies on a wet stool mass basis and did not report the percent solids for their samples, we made assumptions about solids content to directly compare our quantitative data (see Figures S6 and S8 in SI). Overall, the SARS-CoV-2 quantities in our study are higher than those reported in the Wölfel study (Figure S8). Specifically, the Wölfel et al. maximum shedding value of 2.35×10^5^ gc/mg-dw was an order of magnitude lower than the maximum concentration observed in our sample set (2.79×10^6^ gc/mg-dw). Likewise, the geometric mean from Wölfel et al. (157 gc/mg-dw) was two orders of magnitude lower than our geometric mean (1.56×10^4^ gc/mg-dw). These large differences in peak and mean concentrations may be due to the larger number of individuals included in our study (48) compared to the Wölfel study (9).

Interestingly, some of the stool samples measured in our study that were positive for SARS-CoV-2 RNA at 25+ days ASO had relatively high concentrations (i.e. greater than 10^5^ gc/mg-dw). Although the quantities reported in the Natarajan study cannot be directly compared with other studies due to missing data on stool masses, the relative abundance between samples suggests that the maximum fecal concentrations at 25+ days after symptom onset are nearly as high as the maximum concentrations measured at 0-7 days after symptom onset. Combined, the data from our study and the study by Natarajan demonstrate that while fewer infected individuals are shedding SARS-CoV-2 in their feces by 28 days after symptom onset, those that are can be excreting high levels of SARS-CoV-2 RNA.

These data and observations on SARS-CoV-2 fecal shedding have particular value for advancing the field of WBE. For example, the presymptomatic shedding helps explain why wastewater measurements sometimes precede COVID-19 clinical cases.^18^ Likewise, observations of high levels of shedding weeks into an infection suggest that some individuals several weeks into their infection contribute substantially to measured wastewater signals. We anticipate data will be especially impactful for informing mechanistic models. Indeed, the need for high quality fecal shedding datasets has been highlighted in published studies that use wastewater data in epidemiological models.^12–14^ The idiosyncrasies of the shedding trajectories highlight the potential complications of using models to directly predict epidemiological parameters such as disease burden. Such attempts have often relied on static distributions rather than trajectories, however recent work with polio and SARS-CoV-2 have demonstrated methods for incorporating time varying shedding into mechanistic WBE models.^10,30^ Using a static distribution for fecal shedding assumes a uniform likelihood of being at any stage in the shedding period. However, early in an outbreak the majority of individuals are likely in the early stages of infection; while late in an outbreak, it is more likely a mix of early and late stage of infection. We see that different individuals shed at dramatically different rates at different stages of the first 4 weeks after symptom onset. The impact of these patterns remains unknown, but the application of this data in time varying fecal shedding models of WBE systems are necessary to evaluate the impact of biases in a static approach.

This work also fills a critical research gap on the fecal shedding of two commonly used fecal indicator organisms, namely PMMOV and crAssphage. WBE studies routinely present pathogen nucleic acid concentrations on a per PMMoV or crAssphage nucleic acid concentration basis to normalize for differences in wastewater fecal strength and the analytical recovery of viral nucleic acids. Biomarker fecal concentrations have been applied in WBE models,^12,14^ but the data have been limited by the number of individuals observed, the external validity of the measurements, or both. The PMMoV RNA and crAssphage DNA quantities presented here for 48 individuals over time therefore significantly improves the knowledge on the absolute abundance and variability of these biomarkers in stool.

We observed PMMoV in nearly all samples (95.9%) and in at least one sample from all individuals, with a median concentration of 1.00×10^5^ gc/mg-dw and a maximum concentration of 5.0×10^8^ gc/mg-dw. The limited previous studies on PMMoV RNA in stool have detected it less frequently, with 40% of samples from 5 individuals detected by metagenomic techniques^31^ and 66.7% of samples from 9 individuals detected by PCR.^32^ Our PMMoV concentrations are within the range of three stool samples that were quantified in a previous study by RT-qPCR.^32^ That study reported concentrations of 2.3×10^4^, 3.64×10^6^, and 1.95×10^5^ gc/mg-stool. Assuming a 20% dry mass in their samples, the equivalent dry mass concentrations would be 4.6×10^3^, 7.28×10^5^, and 3.90×10^4^ gc/mg-dw, respectively.

CrAssphage DNA was detected in 48.2% of our samples and in at least one sample from 79% of all individuals. The maximum fecal concentration of crAssphage was on the same order of magnitude as the maximum PMMoV fecal concentration, namely 2.41×10^8^ gc/mg-dw; however, the median concentration of crAssphage, 2.13×10^3^ gc/mg-dw, was nearly two orders of magnitude lower than the median concentration of PMMoV. Only one previous study quantified crAssphage shedding in multiple individuals, though the scope of individuals studied was smaller and the issue of external validity remains. Park et al. 2020 measured crAssphage in stool samples from healthy individuals and individuals infected with norovirus.^33^ They observed 70% crAssphage shedding prevalence among two different norovirus outbreaks. This is in contrast with 48% shedding prevalence in healthy adults, and 68.8% shedding in healthy children. Additionally, they observed a wide range in crAssphage concentration, between 10^2.8^ and 10^10.3^ gc/g-stool. It is unclear whether all these samples were from unique individuals, or if certain individuals were sampled multiple times. Other previous studies have documented crAssphage DNA in feces, but they did not provide concentrations.^34–36^

The contrasting distributions of crAssphage and PMMoV shedding could have implications for their use as a normalizing factor for WBE results. It is accepted that the presence of PMMoV RNA in stool is likely due to the consumption of pepper products,^37^ so the variability of PMMoV RNA fecal shedding likely relates to the range of diets between individuals and for individuals over time. These variations may complicate the practice of using it as a normalizing measure of fecal strength. On the other hand, the bimodal fecal concentration distribution of crAssphage suggests that using crAssphage DNA measurements to normalize pathogen measurements may be more biased than PMMoV in some circumstances. Namely, large inconsistencies in the crAssphage concentration could occur depending on which individuals are contributing to the sample. We anticipate this biomarker shedding data will be used in the future in mechanistic mass balance-based models to better evaluate the utility of each biomarker under various sewershed scenarios.

One of the most important aspects of the SARS-CoV-2, PMMoV, and crAssphage datasets presented here is that they are externally-valid. In other words, the laboratory data were collected and reported in a way that makes them useful beyond the context of this study. In this study we achieve external validity in our measurements due to our specific methodologies. For example, we report absolute abundances of SARS-CoV-2 RNA. Relative abundance data, such as those reported as qPCR cycle thresholds (CT) without a standard curve may provide trends within a single study but cannot be reliably compared and combined with data from other studies. Likewise, we measured precise sample mass in our extractions and report our data on a per mass basis. Gene copy data reported per PCR reaction or nucleic acid extract volume without sample mass is not generalizable without making assumptions about sample collection. Finally, by reporting our results on a dry mass basis, we improve the ability to accurately estimate the quantity of SARS-CoV-2 RNA that is shed in an event or over a period of time. This is because the dry mass production of feces amongst populations is less variable than the wet mass production of feces.^38^ Consequently, the conversion of SARS-CoV-2 measurements in fecal samples to the SARS-CoV-2 generated in feces by an individual will carry less uncertainty when the fecal data are reported on a per dry mass basis.

We note that the identified limitations of the available SARS-CoV-2 fecal shedding literature is likely due to different priorities between fields. For WBE, accurate fecal shedding data that is reported as absolute abundance per dry fecal sample mass is critical for linking observations in wastewater with population epidemiological measures, such as infection prevalence or R_0_ values. In other applications, SARS-CoV-2 fecal shedding has been investigated to better define COVID-19 disease and help in the identification and treatment of participants. In these cases, measurements that identify the presence/absence of the target or the relative abundance of the target between samples, as opposed to externally valid absolute abundances, may be sufficient. Nonetheless, externally valid and accurate quantitative data will have the broadest utility and benefit a wider range of fields interested in fecal shedding. We therefore encourage future fecal shedding studies to pursue external validity of fecal shedding measurements and incorporate dry mass data.

There are several limitations to our data set as well as opportunities for future work. First, the subjects included in this study were from a relatively small geographic area–all were residents of the San Francisco Bay area. Additionally, the age and demographic distributions, including vaccination status, of the sample population were not large and diverse enough to identify demographic effects on the fecal shedding of SARS-CoV-2, PMMoV, and crAssphage nucleic acids. Furthermore, the samples measured in this study were collected between September 2020 and April 2021, prior to the emergence of the delta or omicron variants and we do not yet know how fecal shedding dynamics are affected by different variants. A final limitation of the biomarker data is that these measurements were only made in individuals who had tested positive for SARS within 30 days. As discussed earlier, there is little data on crAssphage or PMMoV shedding in healthy individuals, as a result the effect of SARS-CoV-2 infection remains unresolved. Despite these limitations, these externally valid quantitative data fill knowledge gaps on SARS-CoV-2 and viral biomarker fecal shedding and are critical for the advancement of the emerging field of WBE. Due to the external validity, our data can be directly compared and consolidated with future SARS-CoV-2 shedding studies, including those focused on the effects of demographics, vaccine status, and variants on fecal shedding.

## Methods

### Cohort Description and Stool Sample Collection

Stool samples were collected by the FIND COVID project, a CDC-funded cohort study of SARS-CoV-2 infectivity and viral transmission among households in the San Francisco Bay area.^39,40^ Individuals with positive, provider ordered PCR tests for SARS-CoV-2 RNA were eligible if they were not hospitalized and lived in close contact with at least one other individual. Household contacts of index cases were defined as anyone who spent at least one night in the household any time between two days before illness onset, through index case enrollment and tested positive for SARS-CoV-2 after the exposure. Household contacts were ineligible if they had any previous history of SARS-CoV-2 infection or had suspected infection in the 14 days leading up to index case illness onset. Index cases and their household members were instructed to self-collect stool samples periodically over approximately 30 days after index case symptom onset. Specimens were collected in 50 mL conical stool sampling tubes before immediate storage at - 20C. Samples were transferred to -80C within one week of collection where they were stored for up to 8 months. Samples were shipped from the University of San Francisco to the University of Michigan on dry ice, where they were immediately stored at -80C prior to extraction and analysis.

### Sample Processing

Protocols for stool sample processing and extraction were adapted from previously published procedures for the analysis of SARS-CoV-2 RNA in fecal samples.^41^ Prior to analysis, stool samples were thawed on ice from -80C. Between 10 and 25 mg of sample was added to a 2 mL screw cap tube with a rubber O-ring (Corning, CAT #430915). Next, 1200 uL of DNA/RNA shield (Zymo, CAT #R1100) and 0.5 g silica zirconia beads (Biospec, CAT #11079105z) were added and the samples were homogenized using a Biospec bead beater (Biospec, CAT # 1001). BCoV vaccine (Merck) was resuspended according to the manufacturer’s instructions and was added to the homogenate as a recovery control (1.5 uL/mL - DNA/RNA shield). Total nucleic acid was extracted with the Chemagic Viral DNA/RNA 300 kit H96 (Perkin Elmer #CMG-1033-S) using the Chemagic 360 automated extraction platform (Perkin Elmer #CMG-360). Samples were extracted in triplicate, along with triplicate samples of nuclease free water as negative extraction controls. Extracts were further processed with Zymo PCR inhibitor removal kits (Zymo, CAT #D6035), following manufacturer instructions. RNA samples were stored at 4 C prior to analysis.

The dry mass fraction for each sample was measured and is included in the supplementary information. In short, sample dry mass fraction was measured by weighing a portion of the sample and placing it in a microcentrifuge tube with a hole pierced in the cap for venting. The tubes were placed in a heating block at 99C for 24 hours before the sample was weighed again.

### ddPCR Analysis

All ddPCR reactions were performed using the Advanced One-step RT-ddPCR Advanced kit for Probes (Biorad, Cat# 1864022). Two different ddPCR approaches were used to quantify the targets of interest. At first, a duplex RT-ddPCR assay was used and RNA extracts were analyzed twice, once for SARS-CoV-2 N and ORF1a, and again for BCoV and PMMoV. This approach used separate channels for each target using the following concentrations of primers and probes: 2.7 uM of each primer and 0.75 uM of each probe. Later, a multiplexed RT-ddPCR assay to quantify all four targets in a single plate was implemented to be more resource efficient. Such a technique is described in the dMIQE guidelines^26^ and was validated in our lab to produce comparable results from the same samples (figure S9). Primers and probes used in these assays were developed previously and have been used in published research (Table S1).^4^ In short, 22 uL reactions contained 1.8 uM of SARS-CoV-2 N and ORF1a gene primers, 0.5 uM N (FAM) and ORF1a (HEX) hydrolysis probes, 0.9 uM PMMoV and BCoV primers, and 0.25 uM PMMoV (HEX) and BCoV (FAM) probes. All other reaction components were included at concentrations suggested by the manufacturer. Triplicate extraction replicates were plated for each sample, along with triplicate extraction negative controls and triplicate no template controls. Triplicate mixed positive controls containing all four targets: (1) PMMoV gBlock gene fragment (IDT), (2) extracted RNA from the BCoV vaccine, and (3) SARS-CoV-2 gene fragment (NIST), were run on each plate. For each sample, extract dilutions of 1:10 and 1:100 were included to ensure optimal quantification and to identify inhibition effects. Analysis was performed using a BioRad ddPCR system (Biorad, Cat #1864100).

Thresholding was conducted with Bio-Rad QuantaSoft Analysis Pro Software (version 1.0). Only wells with > 10,000 total droplets generated were included in the analysis. The limit of blank was calculated from the upper 95% confidence limit of the extraction negative control on each reaction plate.^29^ Reaction concentrations (gene copies/reaction) given by the instrument were converted to gene copies/mg-dry weight using the mass of the sample extracted and the dry mass fraction of the sample, a sample calculation is provided in the supplementary information (Equation S1). RT-ddPCR inhibition was observed in many of the measured samples, where the 1:100 extract dilutions ultimately resulted in higher target concentrations than the 1:10 dilutions. The influence of PCR inhibition was minimized by adopting the following procedure: when multiple dilutions of a sample provided a measurement above the detection limit, the highest dilution was selected. If none of the dilutions provided a measurement above the LOB, the LOB of the least diluted replicate was recorded as a non-detect . BCoV was measured as a positive recovery control, and to ensure amplification and validate dilution series. Data was not used from samples where the BCoV recovery was less than 50%.

### qPCR Analysis

A separate qPCR assay was used to quantify crAssphage DNA as adding in a 5th target for the ddPCR made thresholding unreliable. Additionally, qPCR has a much larger single-reaction dynamic range which proved useful for the highly variable crAssphage DNA concentrations in stool. Using a qPCR approach eliminated the need of plating multiple dilutions per sample. The primers and probe used in this study were developed previously.^42^ Additional sequence information is included in table S2. PCR reactions were performed using the Luna Universal Probe qPCR master mix (New England Biolabs, Cat# M3004), according to the manufacturer’s instructions. Cycle threshold (Ct) data was converted to gene copies/reaction using a calibration curve consisting of 8 dilutions, in duplicate, of a gBlock gene fragment (IDT) synthesized for the amplicon of interest. Dilutions of the standard were targeted to achieve a dynamic range of 1 gc/uL-rxn to 10^8^ gc/uL-rxn. The gBlock was quantified by duplicate measurements of three different dilutions using a ddPCR assay run concurrently to each qPCR run. The mean dilution corrected concentration was then used to extrapolate across the standard curve. The R^2^ of the linear regression of the standard curve was always greater than .9865 and the amplification efficiency was always greater than 87.4%. Triplicate negative extraction controls and no-template controls were included with every run and were considered acceptable if the concentrations were less than 1.5 cp/uL-rxn. The limit of detection (LOD) was the concentration corresponding to a Ct of 40 (the last cycle of amplification), and the limit of quantification (LOQ) corresponded to the lowest concentration dilution of the standard curve measured on each plate.

### Statistical Analysis

All statistical analyses were performed in R studio (version 4.1.2). To examine dependency of SARS-CoV-2 fecal shedding prevalence to different variables, we used mixed effect logistic regression. Logistic regressions were run using the ‘lme4’ package with the glmer() function with the argument “family = ‘binomial’”.^43^ The formula used in this command was: SARS-CoV-2 shedding (T/F) ∼ Day after symptom onset + Sex + (1 | individual identifier).

To investigate the dependency of the magnitude of crAssphage and PMMoV shedding, we used linear regression. This analysis was performed using the lmer() function from the lmer4 package. In this analysis the formula we used was log10(biomarker_concentration) ∼ Sex + SARS-CoV-2 shedding (T/F) + log10(alternative_biomarker_concentration) + (1 | individual identifier).

## Data Availability

All data produced in the present study are available upon request to the authors

---

**Table 1:** Summary of PMMoV and crAssphage nucleic acid shedding data from the samples measured in this study.

|  | PMMoV (gc/mg-dw) | CrAssphage (gc/mg-dw) |
| --- | --- | --- |
| Geometric Mean | $8.70 \times 10^4$ | $1.44 \times 10^4$ |
| Geometric SD | 37.1 | 148 |
| Median | $1.00 \times 10^5$ | $1.86 \times 10^3$ |
| Fraction of Samples Positive | 0.96 (352/367) | 0.48 (179/371) |
| Fraction of Individuals | 1.00 (48/48) | 0.79 (38/48) |
| Positive |  |  |

