## Supplemental Figures for "Longitudinal and Quantitative Fecal Shedding Dynamics of SARS-CoV-2, Pepper Mild Mottle Virus and CrAssphage"

### Supplementary Information

Table S1: Cohort Demographic Data

| | Number (N = 48) | Percent |
|---|---|---|
| **Sex** | | |
| Male | 21 | 43.75 |
| Female | 27 | 56.25 |
| **Age** | | |
| 1-17 | 4 | 8.3 |
| 19-54 | 36 | 75 |
| 55-70 | 8 | 16.7 |
| **Race/Ethnicity** | | |
| White | 18 | 37.5 |
| Asian | 13 | 27 |
| Hispanic/Latino | 12 | 25 |
| Black/African American | 2 | 4.2 |
| American Indian or Alaska Native | 1 | 2.1 |
| Pacific Islander/Native Hawaiian | 1 | 2.1 |
| Declined to Answer | 1 | 2.1 |

Table S2: Individual target gene ddPCR operating parameters and primer and probe sequences

| Target Gene | Primer Sequence | Cycling Conditions | Amplicon Size | Reference |
|---|---|---|---|---|
| SARS-CoV-2 N | Fw: CATTACGTTTGGTGGACCCT<br>Rv: CCTTGCCATGTTGAGTGAGA<br>Probe: CGCGATCAAAACAACGTCGG 5'FAM/ZEN/3'IBFQ | 50C - 60min<br>95C - 5min<br>[95C - 30s<br>56C - 1min] x40<br>98C - 10min<br>4C - Hold | 143 bp | Wolfe et al. 2021 |

| | | | | |
|---|---|---|---|---|
| SARS-CoV 2 ORF1a | Fw: CAGAACTGGAACCACCTTGT<br>Rv: TACAGTTGAATTGGCAGGCA<br>Probe: TGCCACAGTACGTCTACAAGC<br>5'HEX/ZEN/3'IBFQ | 50C - 60min<br>95C - 5min<br>[95C - 30s<br>56C - 1min] x40<br>98C - 10min<br>4C - Hold | 179 bp | Wolfe et al. 2021 |
| BCoV | Fw: CTGGAAGTTGGTGGAGTT<br>Rv: ATTATCGGCCTAACATACATC<br>Probe: CCTTCATATCTATACACATCAAGTTGTT<br>5'FAM/ZEN/3'IBFQ | 50C - 60min<br>95C - 5min<br>[95C - 30s<br>56C - 1min] x40<br>98C - 10min<br>4C - Hold | 85 bp | Wolfe et al. 2021 |
| PMMoV | Fw: GAGTGGTTTGACCTTAACGTTTGA<br>Rv: TTGTCGGTTGCAATGCAAGT<br>Probe: CCTACCGAAGCAAATG<br>5'HEX/ZEN/3'IBFQ | 50C - 60min<br>95C - 5min<br>[95C - 30s<br>56C - 1min] x40<br>98C - 10min<br>4C - Hold | 68 bp | Wolfe et al. 2021 |
| CrAssphage CPQ064 | Fw: TGTATAGATGCTGCTGCAACTGTACTC<br>Rv: CGTTGTTTTCATCTTTATCTTGTCCAT<br>Probe: CTGAAATTGTTCATAAGCAA<br>5'FAM/ZEN/3'IBFQ | 95C - 5 min<br>[95C - 30s<br>56C - 1min] x40<br>98C - 10min<br>4C - Hold | 126 bp | Stachler et al. 2017 |

Table S3: Estimates derived from logistic regression of SARS-CoV-2 fecal shedding prevalence for an outcome variable of log10(gc/mg-dw)

| **Random Effects** | | | | | |
|---|---|---|---|---|---|
| **Groups** | **Name** | **Variance** | **Std. Dev** | | |
| **ID** | (Intercept) | 13.24 | 3.638 | | |
| **Fixed Effects** | | | | | |
| **Name** | **Type** | **Estimate** | **Std. Error** | **z-value** | **p-value** |
| **Intercept** | | 2.81747 | 0.94748 | 2.974 | 0.00294 |
| **Day after symptom onset** | Continuous | -0.22399 | 0.03611 | -6.204 | 5.52e-10 |
| **Sex = Male** | Binary | 0.14498 | 1.22669 | 0.118 | 0.90592 |

Table S4: Estimates derived from linear regression of concentrations of PMMoV and crAssphage fecal shedding for an outcome variable of log10(gc/mg-dw).

| | | | | | | |
|---|---|---|---|---|---|---|
| **crAssphage** | | | | | | |
| **Random Effects** | | | | | | |
| **Groups** | **Name** | **Variance** | **Std. Dev** | | | |
| **ID** | (Intercept) | 4.6816 | 2.1637 | | | |
| **Residual** | | 0.4219 | 0.6495 | | | |
| **Fixed Effects** | | | | | | |
| **Name** | **Type** | **Estimate** | **Std. Error** | **df** | **t-value** | **p-value** |
| **Intercept** | | 2.70163 | 0.44172 | 55.18068 | 6.116 | 1.03e-07 |
| **Sex = Male** | Binary | -0.51143 | 0.63581 | 45.44786 | -0.804 | 0.425 |
| **Positive SARS-CoV-2 Fecal Shedding** | Binary | 0.09748 | 0.10196 | 318.64308 | 0.956 | 0.340 |
| **log10(PMMoV_conc** | Continuous | 0.02513 | 0.02587 | 312.35886 | 0.971 | 0.332 |
| **PMMoV** | | | | | | |
| **Random Effects** | | | | | | |
| **Groups** | **Name** | **Variance** | **Std. Dev** | | | |
| **ID** | (Intercept) | 0.6787 | 0.8238 | | | |
| **Residual** | | 2.0021 | 1.4149 | | | |
| **Fixed Effects** | | | | | | |
| **Name** | **Type** | **Estimate** | **Std. Error** | **df** | **t-value** | **p-value** |

| | | | | | | |
|---|---|---|---|---|---|---|
| **Intercept** | | 4.78069 | 0.27345 | 64.19427 | 17.483 | <2e-16 |
| **Sex = Male** | Binary | -0.02717 | 0.29561 | 45.30766 | -0.092 | 0.927 |
| **Positive SARS-CoV-2 Fecal Shedding** | Binary | 0.07691 | 0.19406 | 313.24624 | 0.396 | 0.692 |
| **log10(crAss _conc** | Continuous | 0.00974 | 0.06179 | 88.00628 | 0.158 | 0.875 |

Equation S1: Calculation converting the ddPCR generated concentration of gc/uL extract to gc/mg-dw.

$$\frac{Gene\ Copies}{mg\ dry\ weight\ stool} = \frac{Gene\ Copies}{\mu L\ Reaction} \times \frac{22\ \mu L\ Reaction}{5.5\ \mu L\ RNA\ Extract} \times \frac{100\ \mu L\ RNA\ Extract}{300\ \mu L\ Homogenate} \times \frac{1200\ \mu L\ Homogenate}{mg\ Stool} \times \frac{mg\ stool}{mg\ dry\ weight\ stool}$$

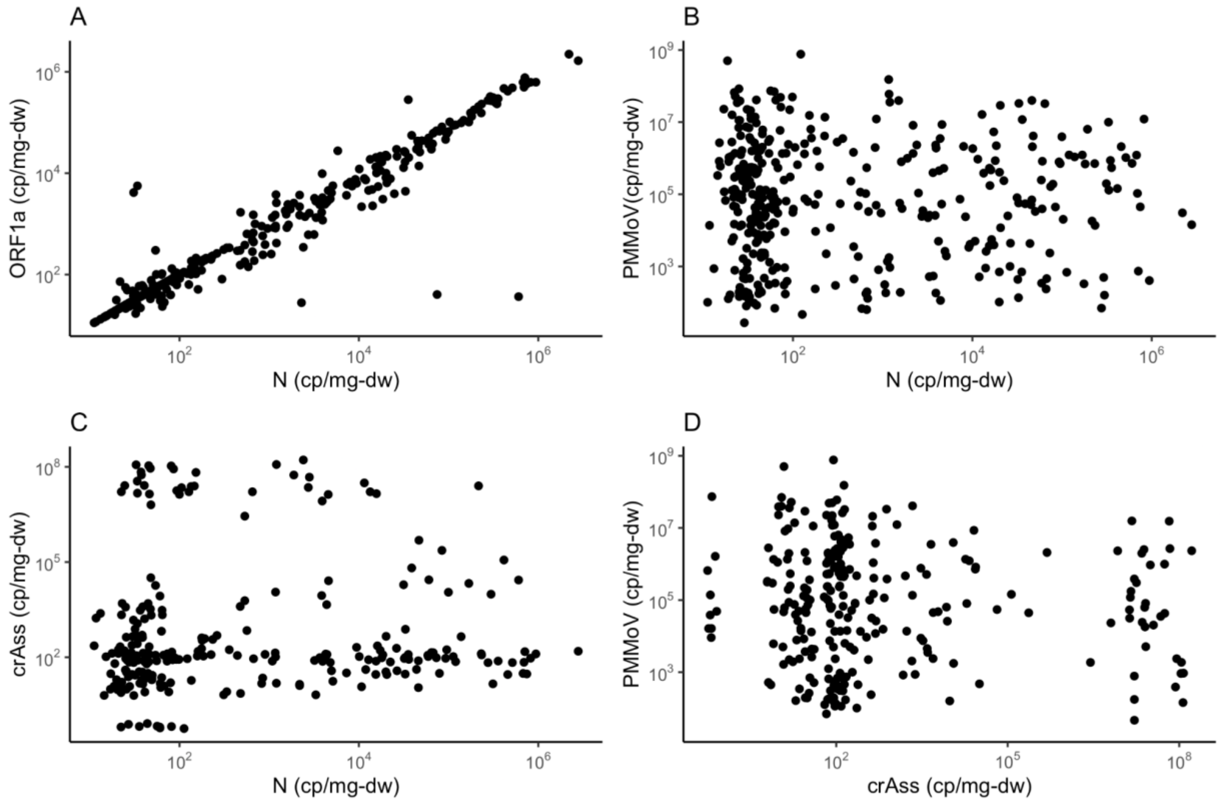

S1. Plots showing correlations between the different gene targets investigated in this study. A) SARS-CoV-2 N vs ORF1a, B) N vs PMMoV, C) N vs. crAss, D) crAss vs PMMoV

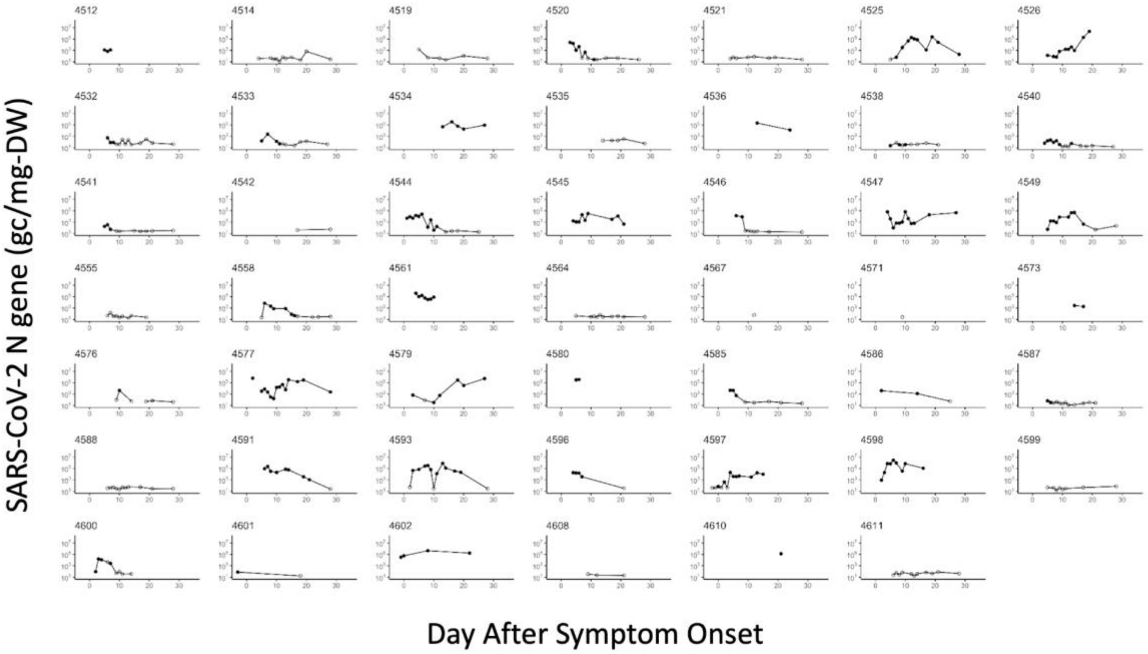

Figure S2. Individual SARS-CoV-2 Trajectories

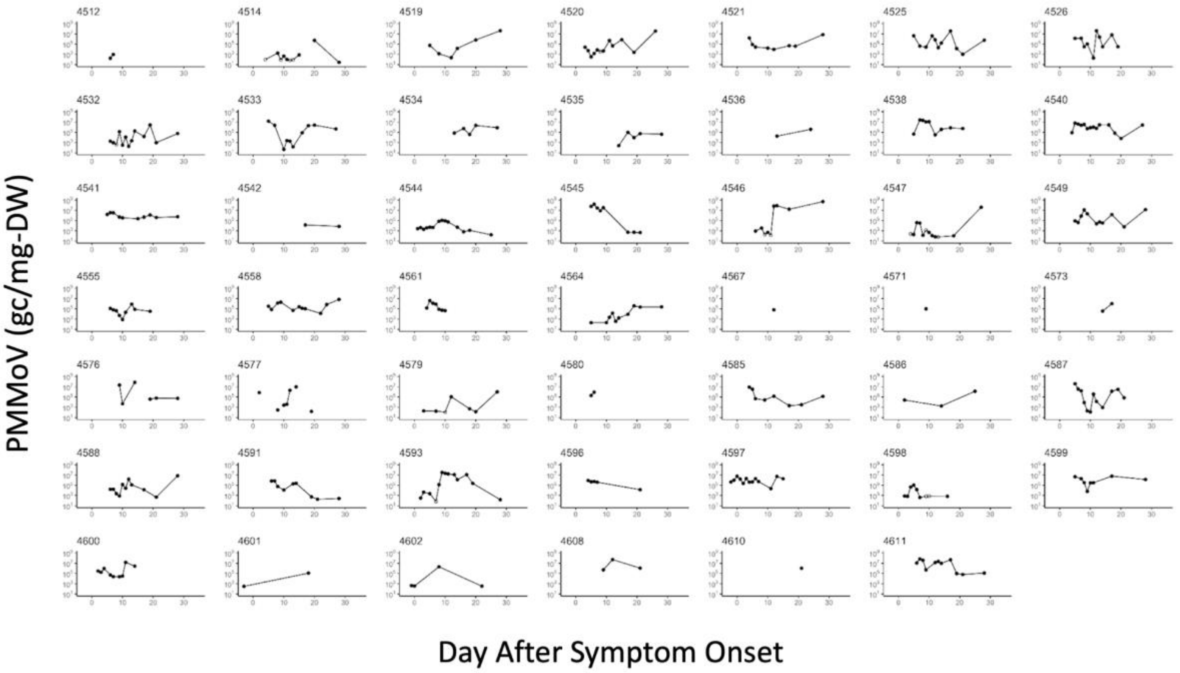

Figure S3: Individual PMMoV shedding trajectories

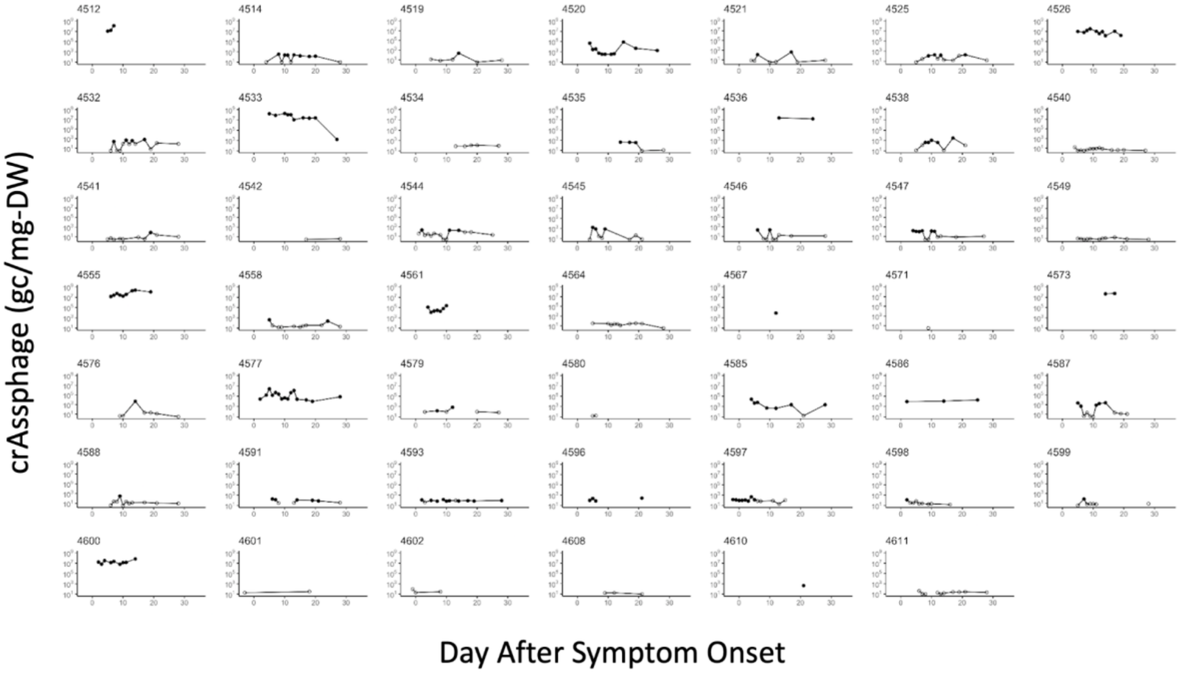

Figure S4. Individual crAssphage shedding trajectories

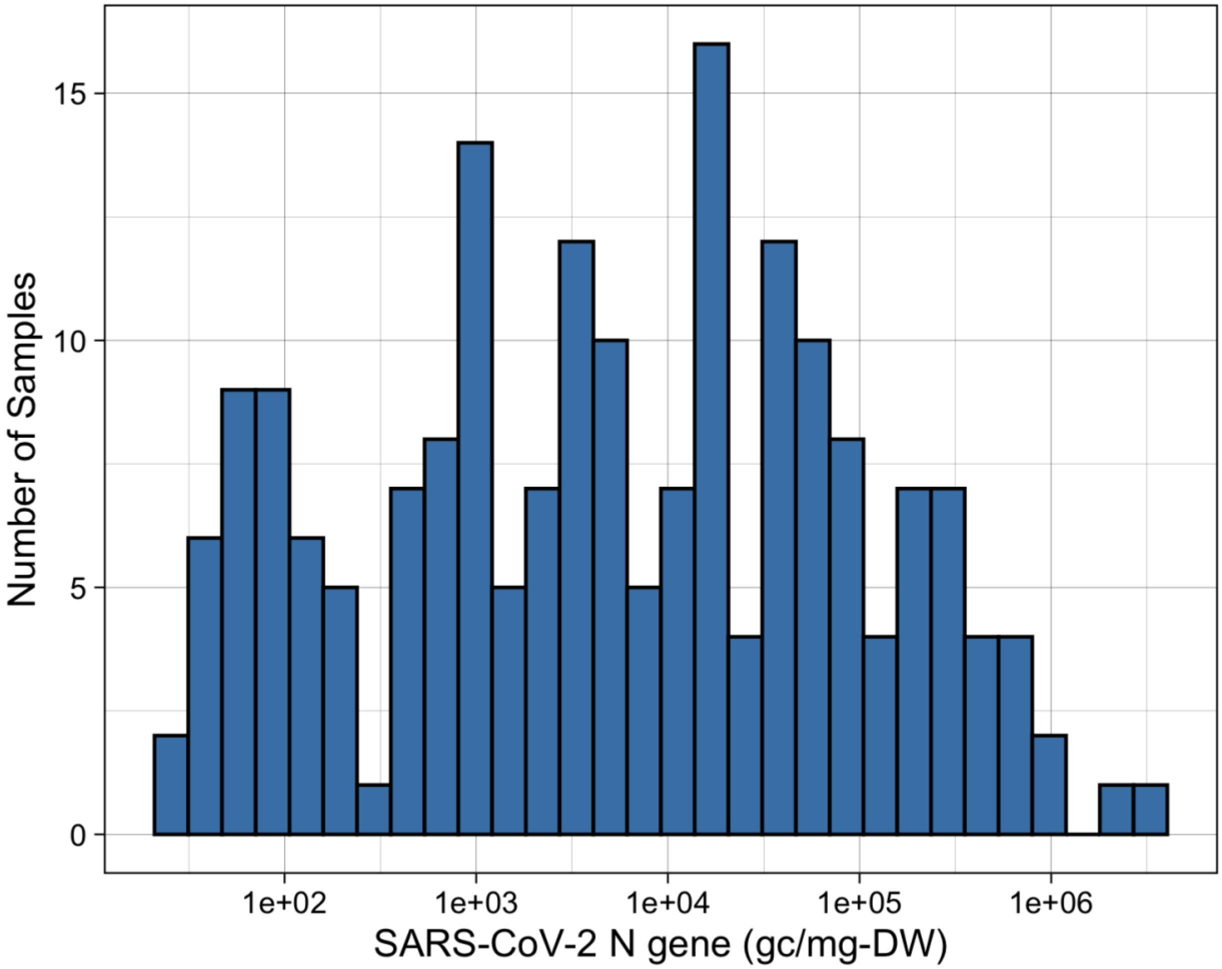

S5: Histogram showing the distribution of all above-LOB SARS-CoV-2 nucleocapsid gene measurements.

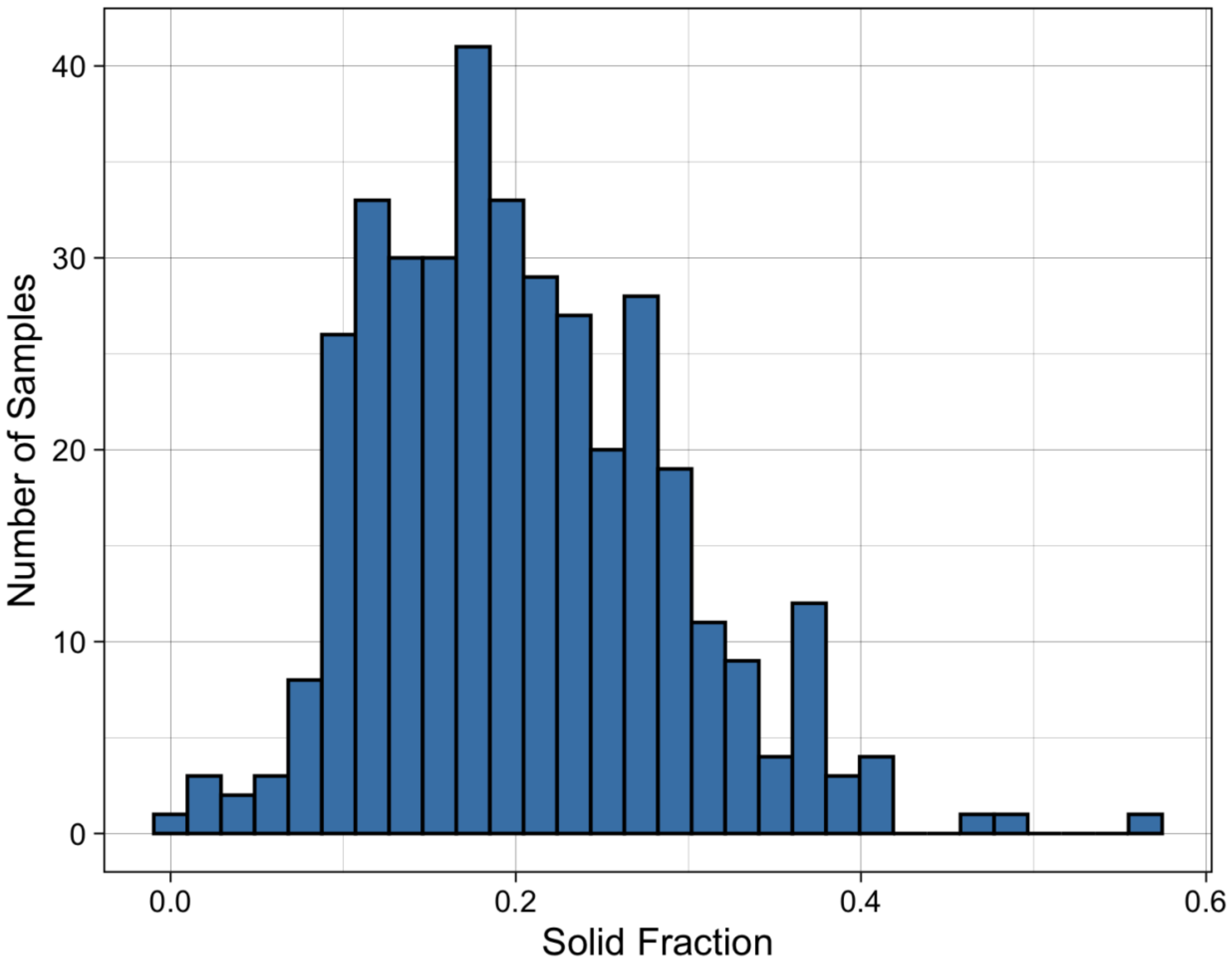

Figure S6: Histogram of the distribution of the solid fraction of all stool samples measured.

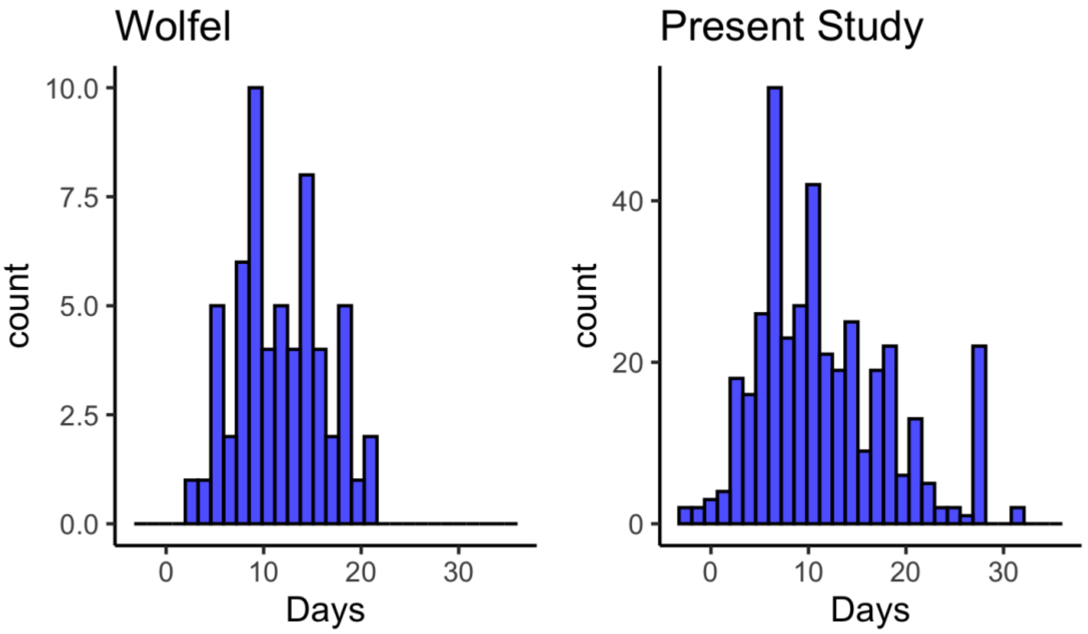

.
Figure S7: Histogram comparing the sampling coverage (post symptom onset) between Wölfel et al 2020, and the present study.

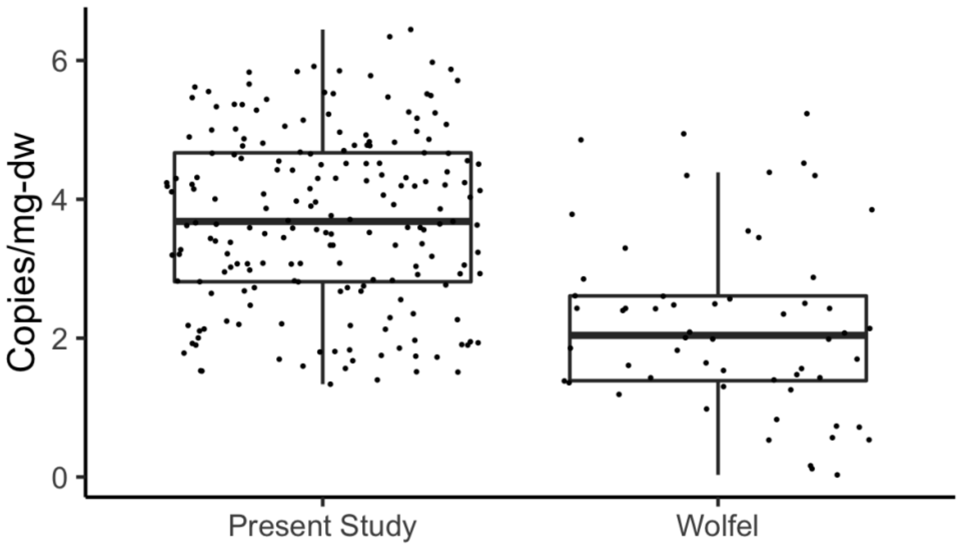

Figure S8: Comparison of the magnitudes of all positive measurements collected from this study with those collected in Wölfel et al 2020, assuming 20% dry mass samples.

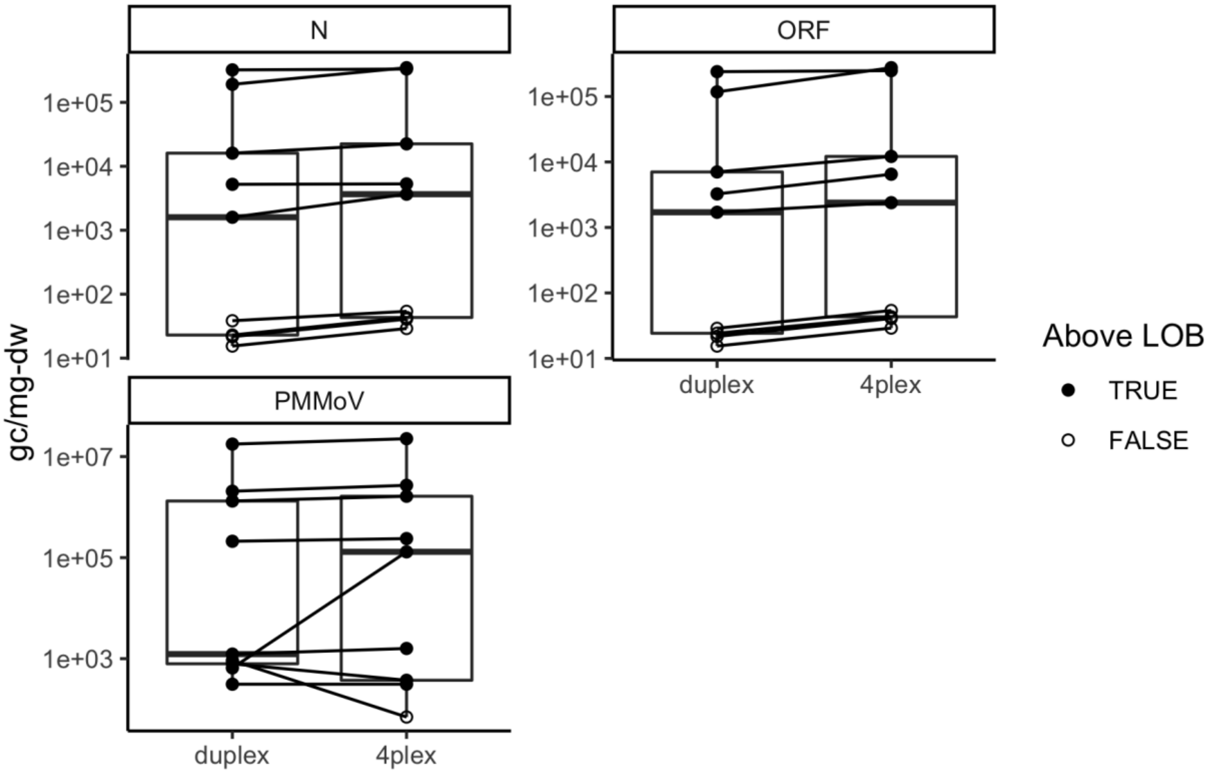

Figure S9: Side-by-side comparison of the RT-ddPCR 4plex assay with two duplex assays for the targets of interest.

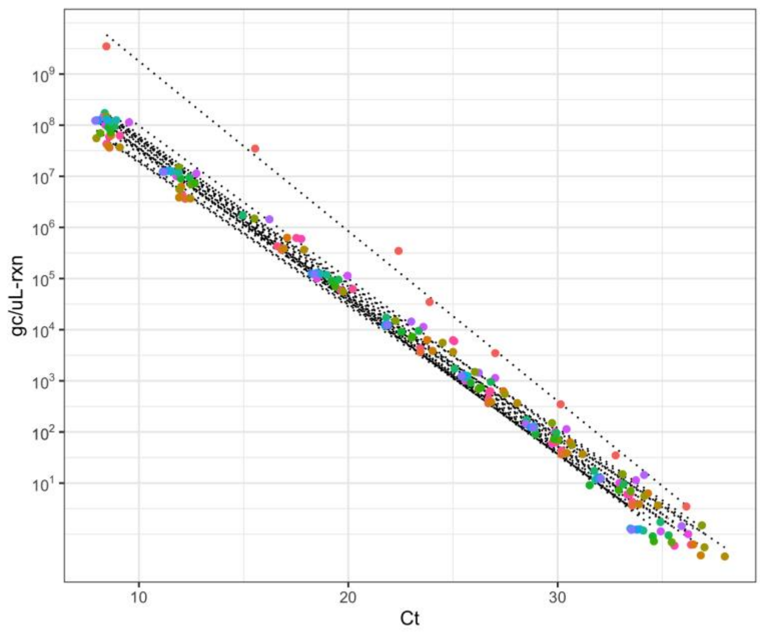

Figure S10: Standard curves correlating the Ct of crAssphage gene fragment dilutions as measured by qPCR to gene copies per microliter of reaction, measured by ddPCR.

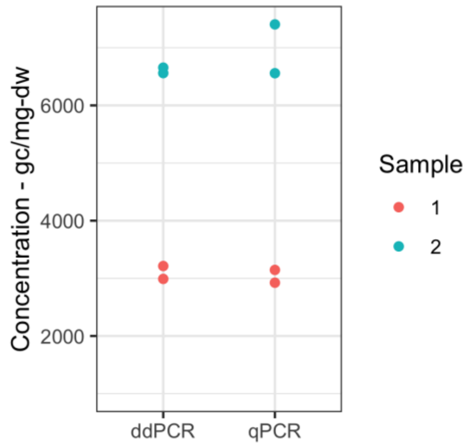

Figure S11: Comparison of quantified concentration of crAssphage positive stool samples from ddPCR vs qPCR with a ddPCR quantified standard curve. For each sample, extraction duplicates were measured using each method.

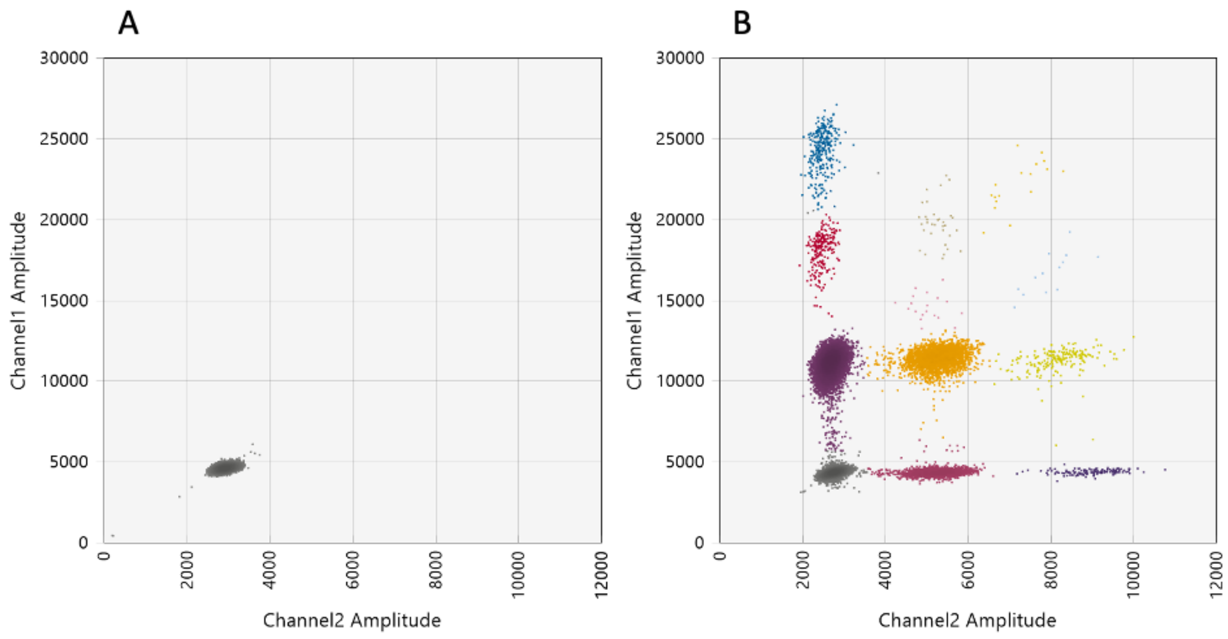

Figure S12: Examples of negative (A), and positive (B) samples measured by the 4 multiplexed ddPCR assay. The positive example exhibits a sample that was positive for all 4 targets.
